# Urinary Biomarkers and Attainment of Cefepime Therapeutic Targets in Critically Ill Children

**DOI:** 10.1101/2021.12.30.21268328

**Authors:** Kevin J. Downes, Anna Sharova, Lauren Gianchetti, Adam S. Himebauch, Julie C. Fitzgerald, Athena F. Zuppa

## Abstract

**INTRODUCTION:** The recommended therapeutic target for cefepime (FEP) is the time above MIC (*f*T>MIC). The frequency of target attainment and risk factors for sub-therapeutic concentrations in children have not been extensively studied.

**METHODS:** We performed a prospective observational pilot study in children in our PICU receiving standard dosing of FEP for suspected sepsis (≥2 SIRS criteria). Three FEP concentrations were measured per subject and a urine sample was collected prior to PK sampling for measurement of urinary biomarkers. We used log linear regression to calculate the *f*T>MIC for each subject across a range of MIC values (1-16 µg/mL). We compared clinical factors/biomarkers between patients who did and did not achieve 100% *f*T>MIC for 8 µg/mL (cut-point for *Pseudomonas*) and tested the correlation between covariates and FEP troughs.

**RESULTS:** 21 subjects were enrolled (median SIRS criteria: 3). PK sampling occurred after a median of 5 doses (range: 3-9). 43% of subjects achieved 100% *f*T>MIC for an MIC of 8 µg/mL. Younger age (p=.005), higher estimated GFR (p=.03), and lower urinary NGAL (p=.006) and KIM-1 (.03) were associated with failure to attain 100% *f*T>8 µg/mL. Age (r = 0.53), eGFR (r = - 0.58), urinary NGAL (r = 0.42) and KIM-1 (r = 0.50) were significantly correlated with FEP troughs.

**CONCLUSIONS:** A significant proportion of critically ill children failed to attain target concentrations for treatment of *Pseudomonas aeruginosa* with FEP. Younger patients and those with good kidney function (high GFR, low urinary biomarkers) may be at highest risk for subtherapeutic FEP concentrations.

## Introduction

Critical illness alters volume of distribution, protein binding and drug clearance, affecting the pharmacokinetics and pharmacodynamics (PK/PD) of antibiotics.^1,2^ β-lactam exposures in critically ill adults are often suboptimal, contributing to poor outcomes.^3,4^ Standard dosing is often also inadequate in critically ill children,^5,6^ especially for treatment of more resistant Gram-negative pathogens.^7^ Pediatric severe sepsis and septic shock have an in-hospital mortality of roughly 6-26%,^8-10^ with delayed antimicrobial administration being associated with increased mortality and organ dysfunction.^10,11^ Thus, early and optimal antibiotic exposures in pediatric sepsis may provide a means to improve patient outcomes.

Cefepime (FEP), a renally excreted 4^th^-generation cephalosporin, is used as empiric antibiotic coverage in many critically ill children with suspected sepsis. Efficacy of FEP is defined by the fraction of time of the dosing interval for which the free (unbound) concentration is maintained above the MIC (*f*T>MIC) of the infecting pathogen.^12^ While the optimal *f*T>MIC associated with clinical efficacy in critically ill children is unknown, the recommended therapeutic target for critically ill adults of *f*T>MIC of 100%.^13^ As such, this target is readily extrapolated to children. Meanwhile, microbiological cure and suppression of resistance selection have been associated with a more robust PK/PD target of C_min_ > 4x MIC,^14^ which equates to 100% *f*T>4x MIC. There is a significant gap in knowledge of the FEP exposures achieved in critically ill children as no studies have reported the frequency of FEP PK/PD target attainment in this vulnerable population.

One of the challenges to optimizing β-lactam therapy in children is knowing who is at greatest risk for sub-therapeutic exposures with standard dosing. Traditional methods for estimating renal function and drug clearance in children using serum creatinine (SCr)-based equations^15^ do not readily identify children with augmented renal clearance (ARC) or changing glomerular filtration in the setting of acute kidney injury (AKI). Novel urinary biomarkers, such as neutrophil gelatinase-associated lipocalin (NGAL) and kidney injury molecule-1 (KIM-1), can detect AKI earlier and correlate better with glomerular filtration rate (GFR) and drug clearance than SCr.^16-18^ Thus, they could identify patients with impaired renal clearance at risk for supra-therapeutic drug concentrations. Conversely, low biomarkers levels, signifying the absence of kidney injury, may serve as noninvasive screening tests to detect children at highest risk for ARC and sub-therapeutic drug exposures.

This study sought to test the hypothesis that lower urinary biomarker concentrations (good renal function) were correlated with failure to attain therapeutic targets early in the course of FEP therapy in critically ill children with suspected sepsis due to augmented CL of the drug.

## Methods

### Subjects and Setting

We performed a prospective observational pilot study in children (<19 years of age) admitted to the pediatric intensive care unit (PICU) of the Children’s Hospital of Philadelphia. Children were eligible to participate if prescribed standard dosing of FEP (50 mg/kg/dose every 8 hours if <40 kg; 2000 mg/dose every 8 hours if ≥40 kg) and had ≥2 systemic inflammatory response syndrome (SIRS) criteria present within 48 hours prior to initiation of FEP to be eligible;^19^ SIRS criteria are described in the Supplemental Materials. Children receiving extracorporeal membrane oxygenation, renal replacement therapy, or plasmapheresis, as well those unable to provide urine and blood samples, were excluded. The Institutional Review Board (IRB) approved the study protocol (IRB # 17-014486) with a waiver of documented assent; verbal assent was obtained, as appropriate. Written informed consent was obtained from the subject or parents/legal guardians, as appropriate.

### Cefepime PK sampling

Following enrollment and administration of at least 3 FEP doses, 3 blood samples were collected during a single dosing interval. Samples were collected using lithium-heparin tubes at 3 hours (+/-60 min.), 4.5 hours (+/-60 min), and 6 hours (+/-60 min) after completion of a dose. All sampling was completed by 72 hours of therapy in order to reflect drug concentrations early in FEP therapy; subjects had to provide at least 2 blood samples to be considered evaluable.

Each blood sample was promptly centrifuged (3800 RPM for 5 minutes) following collection, and the plasma portion was aliquoted and stored at -80° C. Total FEP concentrations in plasma were determined in the Bioanalytical Core Laboratory of the Center for Clinical Pharmacology using high-performance liquid chromatography and tandem mass spectrometry, as previously described.^20^ The lower limit of quantification for the assay was 5 ng/mL, with a range of 5-10,000 ng/mL.

### Urinary biomarkers

Prior to blood sampling, a single urine sample was collected for measurement of urinary biomarkers. Samples could be collected at any time prior to FEP blood concentration measurement. Fresh urine was collected from the subject using an indwelling urinary catheter, clean intermittent catheterization, or urine bag, as appropriate. Samples were kept on ice or refrigerated at 4°C until centrifugation (3000 RPM at 4°C for 15 minutes). The supernatants were then divided and stored at -80°C. Urine biomarkers were measured at the CHOP Translational Core Lab via Quantkine® ELISA (NGAL, KIM-1, osteopontin, cystatin C; R&D Systems, Inc., Minneapolis, MN) and the Luminex platform (clusterin; R&D Systems, Inc., Minneapolis, MN). Biomarker concentrations that were out of the range of the assay were assigned a value at the maximum or minimum of the assay range, as appropriate; assay ranges are provided in Supplemental Materials. Urine creatinine (UCr) was also measured on each sample by 2-point end enzymatic method (Roche Diagnostics, Indianapolis, IN). Serum creatinine (SCr) was collected per standard of care and measured in the CHOP Chemistry Laboratory using the 2-point rate spectrophotometric method (Vitros5600™ analyzer, Ortho Clinical Diagnostics, Markham, ON); SCr was rounded to 0.1 if reported as <0.2 mg/dL, which was the limit of quantification for the assay during the study period.

### Data Analysis

Using the measured total FEP concentration, we determined the free FEP concentration assuming protein binding of FEP of 39% (i.e. free FEP = 61% total FEP), based on data from Al-Shaer and colleagues.^21^ We then used log linear regression of free FEP concentrations to estimate the FEP C_min_ and calculate the *f*T>MIC for each subject across a range of MIC values. For this analysis, calculated free concentrations were log-transformed and linear regression used to generate an individual regression line for each subject. From this, we calculated the C_min_ at 8 hours following the dose preceding FEP concentration sampling, as well as the fT>MIC for MIC values of 1, 2, 4, 8, and 16 µg/mL. The proportion of subjects who attained 100% fT>MIC and 100% fT>4x MIC for each of these MIC values was determined. A sensitivity analysis was also performed, using 20% protein binding (80 % free), which has traditionally been used in pediatric studies,^22^ and the above procedures repeated.

Estimated glomerular filtration rate (eGFR) was determined for each subject at FEP initiation and at PK sampling using the bedside Schwartz equation.^23^ Augmented renal clearance (ARC) was defined as an eGFR >130 mL/min/1.73 m^2^, which is the most common cut-point used.^24^ We further dichotomized patients based on an eGFR < 60 mL/min/1.73 m^2^ (renal impairment), as an exploratory analysis.

We tested the association between covariates (clinical characteristics, biomarkers) and 100% *f*T>MIC for an MIC of 8 µg/mL, the CLSI breakpoint for *Pseudomonas aeruginosa*, using chi-squared and Wilcoxon rank sum tests; this cut-point was selected since *Pseudomonas* is an organism of high concern that is often targeted with empiric antimicrobial therapy in the critical care setting. We also determined the Spearman correlation between covariates and calculated FEP C_min_ at 8 hours (i.e. trough concentrations). Because of our small sample size and non-normal distribution of continuous variables (e.g. biomarker concentrations), non-parametric tests were utilized for all comparative analyses. We additionally performed a subset analysis among subjects <40 kg (i.e. those who received FEP at a dosage of 50 mg/kg/dose rather than 2000 mg/dose).

All data analyses were conducted using R version 3.6.3 (Vienna, Austria). No formal samples size calculations were made *a priori* since this was an unfunded pilot study designed to explore the potential correlation between FEP concentrations and biomarker concentrations and there were no preexisting data to guide sample size estimations.

## Results

In total 31 subjects were consented to participate in the study from May 2018 through December 2019. Two subjects became ineligible following consent (one was started on plasmapheresis and the other transferred out of the PICU). Eight additional subjects were withdrawn from the study due to inadequate venous access at the time of scheduled FEP sampling. As a result, 21 subjects fully participated in the study and provided adequate urine and blood samples for analysis.

The characteristics of the study population are shown in **Table 1**. The median number of SIRS criteria met upon initiation of FEP was 3: 6 subjects met 2 criteria, 7 met 3, and 8 met all 4. Eighteen subjects had a primary indication for FEP of suspected sepsis, 2 of fever with a central line, and 1 of neutropenic fever. Ten subjects were clinically diagnosed with a bacterial infection and received a definitive course of antibiotic treatment, including 6 with a lower respiratory tract infection, 3 with bacteremia, and 1 with a urinary tract infection; the remainder received FEP empirically. Six subjects had gram-negative bacteria isolated on culture during their clinical evaluation for infection: *Serratia marcescens* (blood, FEP MIC ≤ 1 µg/mL), *Escherichia coli* (blood, FEP MIC ≤ 1 µg/mL), *Pseudomonas aeruginosa* (3 subjects, all respiratory tract isolates; FEP MIC ≤1, 2 and 16 µg/mL), and *Providencia stuartii* (urine, FEP MIC 1 µg/mL). Two subjects had MRSA isolated from a respiratory tract culture, one had *Enterococcus faecalis* isolated from a urine culture, one had MSSA bacteremia, and another had MSSA isolated from a respiratory tract culture. No subjects died during FEP treatment or as a complication of their infection.

**Table 1.**
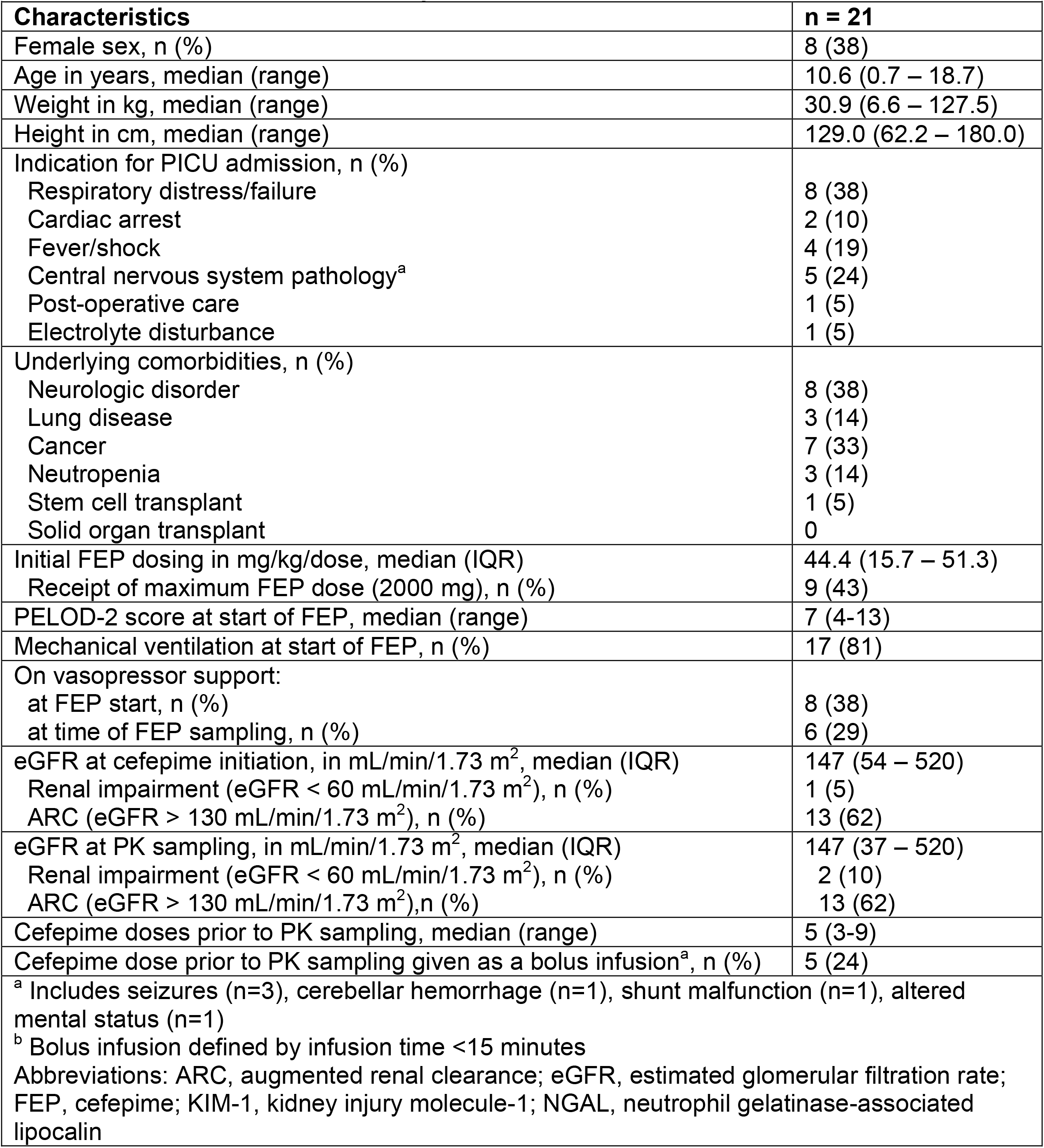
Characteristics of the study population

| **Table 1. Characteristics of the study population.** |  |
| --- | --- |
| **Characteristics** | **n = 21** |
| Female sex, n (%) | 8 (38) |
| Age in years, median (range) | 10.6 (0.7 – 18.7) |
| Weight in kg, median (range) | 30.9 (6.6 – 127.5) |
| Height in cm, median (range) | 129.0 (62.2 – 180.0) |
| Indication for PICU admission, n (%) |  |
| Respiratory distress/failure | 8 (38) |
| Cardiac arrest | 2 (10) |
| Fever/shock | 4 (19) |
| Central nervous system pathology<sup>a</sup> | 5 (24) |
| Post-operative care | 1 (5) |
| Electrolyte disturbance | 1 (5) |
| Underlying comorbidities, n (%) |  |
| Neurologic disorder | 8 (38) |
| Lung disease | 3 (14) |
| Cancer | 7 (33) |
| Neutropenia | 3 (14) |
| Stem cell transplant | 1 (5) |
| Solid organ transplant | 0 |
| Initial FEP dosing in mg/kg/dose, median (IQR) | 44.4 (15.7 – 51.3) |
| Receipt of maximum FEP dose (2000 mg), n (%) | 9 (43) |
| PELOD-2 score at start of FEP, median (range) | 7 (4-13) |
| Mechanical ventilation at start of FEP, n (%) | 17 (81) |
| On vasopressor support: |  |
| at FEP start, n (%) | 8 (38) |
| at time of FEP sampling, n (%) | 6 (29) |
| eGFR at cefepime initiation, in mL/min/1.73 m<sup>2</sup>, median (IQR) | 147 (54 – 520) |
| Renal impairment (eGFR < 60 mL/min/1.73 m<sup>2</sup>), n (%) | 1 (5) |
| ARC (eGFR > 130 mL/min/1.73 m<sup>2</sup>), n (%) | 13 (62) |
| eGFR at PK sampling, in mL/min/1.73 m<sup>2</sup>, median (IQR) | 147 (37 – 520) |
| Renal impairment (eGFR < 60 mL/min/1.73 m<sup>2</sup>), n (%) | 2 (10) |
| ARC (eGFR > 130 mL/min/1.73 m<sup>2</sup>), n (%) | 13 (62) |
| Cefepime doses prior to PK sampling, median (range) | 5 (3-9) |
| Cefepime dose prior to PK sampling given as a bolus infusion<sup>a</sup>, n (%) | 5 (24) |
| <sup>a</sup> Includes seizures (n=3), cerebellar hemorrhage (n=1), shunt malfunction (n=1), altered mental status (n=1) |  |
| <sup>b</sup> Bolus infusion defined by infusion time <15 minutes |  |
| Abbreviations: ARC, augmented renal clearance; eGFR, estimated glomerular filtration rate; FEP, cefepime; KIM-1, kidney injury molecule-1; NGAL, neutrophil gelatinase-associated lipocalin |  |

Nine subjects received the maximum FEP dosage of 2000 mg/dose. Thirteen subjects (62%) had ARC at the time of PK sampling. Blood sampling for FEP concentration measurement occurred following a median of 5 doses (range 3-9). The median time of urine sample collection following FEP start was 25.8 hours (IQR: 22.9 – 38.5 hours); median time from urine sample collection to first PK measurement was 8.9 hours (IQR: 1.0 – 15.6 hours). Three subjects had clusterin concentrations that were below the limit of quantification (BQL) of the assay and 3 had KIM-1 concentrations below that were BQL. Two and one subjects had NGAL and osteopontin concentrations above the upper limits of the assay ranges, respectively.

Based on log-linear regression, the median estimated FEP C_min_ was 4.61 µg/mL (range: 0.63 – 62.39 µg/mL). **Table 2** displays the fraction of subjects with C_min_ ≥ MIC and C_min_ ≥ 4x MIC for MICs from 1-16 µg/mL. Ninety percent of subjects achieved 100% *f*T>MIC for an MIC of 2 µg/mL, decreasing to 52% for an MIC of 4 µg/mL, 43% for an MIC of 8 µg/mL, and 33% for an MIC of 16 µg/mL. **Figure 1** displays *f*T>MIC for the study population based on varying MIC targets. This figure was recreated assuming 20% protein binding and is shown in Supplemental Materials. The number of subjects attaining a C_min_ ≥ 8 µg/mL were the same when protein binding of 20% and 39% were used.

**Table 2.** Target attainment for subjects treated with cefepime^a^

| **Table 2. Target attainment for subjects treated with cefepime<sup>a</sup>** |  |  |
| --- | --- | --- |
| MIC (µg/mL) | Cmin ≥ MIC, n (%) | Cmin ≥ 4xMIC, n (%) |
| 1 | 20 (95.2) | 11 (52.3) |
| 2 | 19 (90.4) | 9 (42.9) |
| 4 | 11 (52.3) | 7 (33.3) |
| 8 | 9 (42.9) | 4 (19.0) |
| 16 | 7 (33.3) | 0 |
| <sup>a</sup> Target attainment determined by performance of log-linear regression of unbound cefepime concentrations on time. Plasma protein binding assumed to be 39% for all calculations (based on data by Al-Shaer MH, et al. *Ther Drug Monit*, 2020). |  |  |

**Figure 1.**
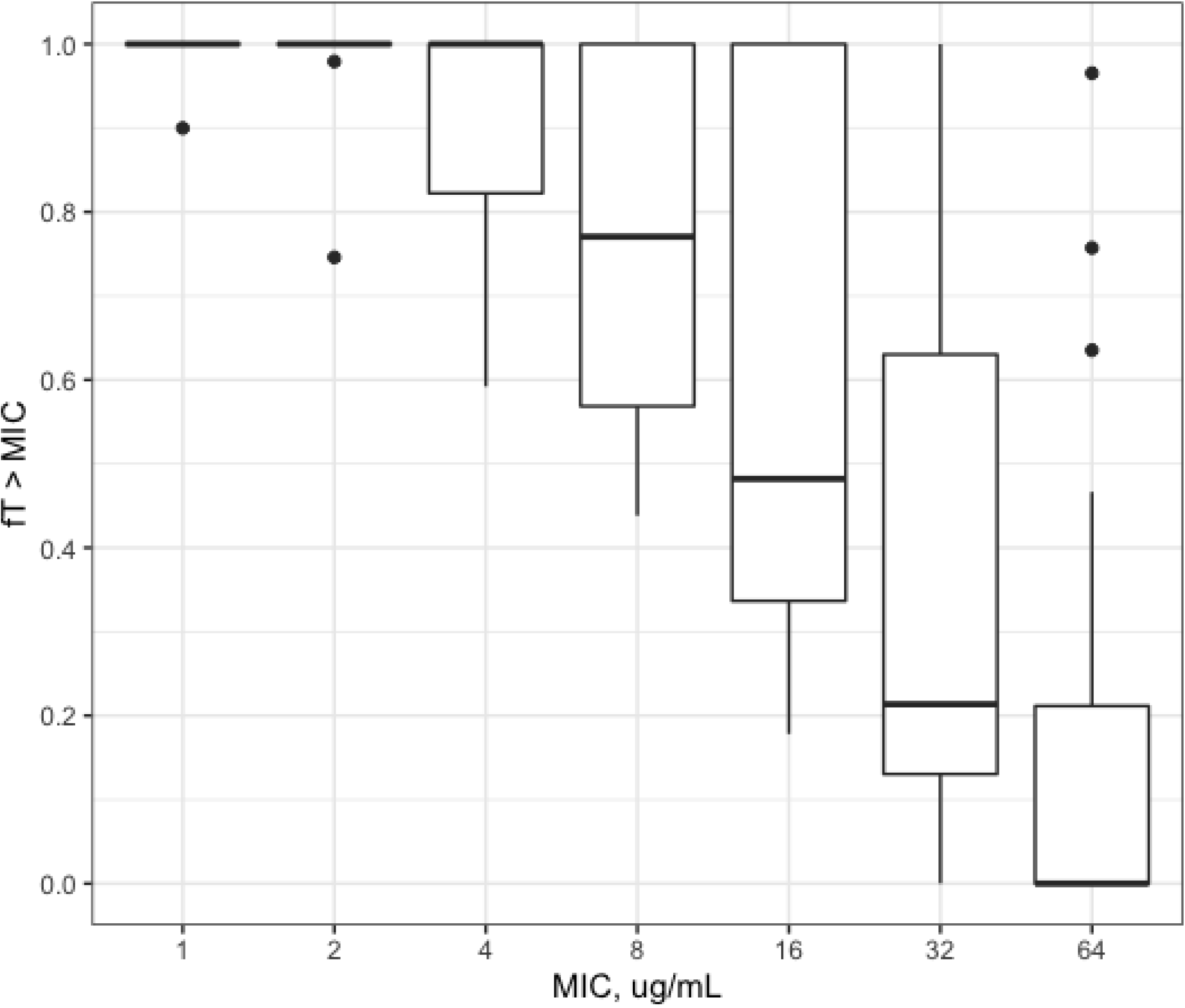
Box-and-whisker plots displaying the fraction of the dosing interval that free FEP concentrations remained above the MIC among study subjects. The free fraction of FEP was calculated based on 39% protein binding.^21^ For each plot, the thick solid line represents the median, the box displays the inter-quartile range (IRQ, 25^th^-75^th^ percentile), whiskers incorporate 1.5 times the inter-quartile range, and the dots are values outside of 1.5 times the IQR.

Twelve subjects (57%) had a FEP C_min_ below 8 µg/mL (i.e. failed to meet the target of 100% *f*T>MIC for an MIC of 8 µg/mL). These subjects were younger and had a higher eGFR at the time of FEP measurement (**Table 3**). Subjects with ARC more often had a C_min_ below 8 µg/mL (9/13 [69%] if eGFR >130 mL/min/1.73 m^2^ vs 3/8 [38%] with eGFR <130), although this difference was not statistically significant (p = 0.20). Subjects with FEP C_min_ <8 µg/mL also had lower NGAL and KIM-1 urinary concentrations (both as unadjusted concentrations and when biomarker concentrations were adjusted for urinary creatinine). Higher age and lower eGFR at the time of FEP sampling were significantly correlated with FEP C_min_ (**Table 4**). Urinary NGAL and KIM-1, as well as urinary NGAL/UCr and KIM-1/UCr, were also significantly correlated with FEP C_min_.

**Table 3.** Comparison between clinical factors and cefepime C_min_ > 8 µg/mL

| **Table 3.** Comparison between clinical factors and cefepime $C_{\min} > 8 \mu\text{g/mL}$ | | | |
| --- | --- | --- | --- |
| **Factor** | **$C_{\min} < 8$**<br>**(n = 12)** | **$C_{\min} \geq 8$**<br>**(n = 9)** | **p-value** |
| Age in years, median | 2.6 | 16.6 | **.005** |
| Weight in kg, median | 16.3 | 45 | .11 |
| Dosage in mg/kg/dose, median | 45.0 | 40.9 | .48 |
| eGFR at FEP sampling, in mL/min/1.73 m<sup>2</sup>, median | 193 | 109 | **.03** |
| ARC at FEP sampling (eGFR > 130 mL/min/1.73 m<sup>2</sup>), % | 75% | 44% | .20 |
| SCr at PK sampling, in mg/dL, median | 0.2 | 0.6 | **.01** |
| PELOD-2 score at start of FEP, median | 7.5 | 7 | .37 |
| Receipt of vasopressors at FEP sampling, % | 25% | 33% | .91 |
| Receipt of FEP as bolus infusion, % | 25% | 22% | 1.0 |
| **Urinary biomarkers, median, ng/mL, median** |  |  |  |
| Cystatin C | 24 | 376 | .11 |
| Clusterin | 51 | 437 | .21 |
| NGAL | 12 | 350 | **.006** |
| Osteopontin | 991 | 1949 | .31 |
| KIM-1 | 0.4 | 1.6 | **.03** |
| **Urinary biomarkers, ng/mg urine creatinine, median** |  |  |  |
| Cystatin C | 141.0 | 863.9 | .08 |
| Clusterin | 204.2 | 814.1 | .19 |
| NGAL | 57.1 | 826.7 | **.004** |
| Osteopontin | 5075.2 | 3728.8 | .65 |
| KIM-1 | 1.9 | 8.9 | **.05** |
| Wilcoxon rank sum tests used for comparisons of continuous variables; Fisher's exact tests used for comparisons of categorical variables. |  |  |  |
| Abbreviations: ARC, augmented renal clearance; $C_{\min}$, minimum concentration; eGFR, estimated glomerular filtration rate; FEP, cefepime; KIM-1, kidney injury molecule-1; NGAL, neutrophil gelatinase-associated lipocalin; PELOD-2, Pediatric Logistic Organ Dysfunction-2; SCr, serum creatinine | | | |

**Table 4.** Spearman correlation analyses between covariates and cefepime C_min_ (i.e. trough).

|  |  |
| --- | --- |
| **Factor** | **r (p-value)** |
| Age | .53 (.01) |
| Weight | .40 (.07) |
| Dose in mg/kg/dose | -.29(.21) |
| eGFR at cefepime initiation | -.32 (.16) |
| eGFR at PK sampling | -.58 (.006) |
| **Urinary biomarkers, ng/mL** |  |
| Cystatin C | .29 (.20) |
| Clusterin | .32 (.15) |
| NGAL | .42 (.056) |
| Osteopontin | .37 (.10) |
| KIM-1 | .50 (.02) |
| **Urinary biomarkers, ng/mg urine creatinine** |  |
| Cystatin C | .34 (.13) |
| Clusterin | .39 (.08) |
| NGAL | .47 (.03) |
| Osteopontin | .18 (.42) |
| KIM-1 | .49 (.02) |

On subset analysis of subjects <40 kg (n= 12), four subjects (33%) had a FEP C_min_ ≥ 8 µg/mL. Older age was associated with attainment of this target, although eGFR was not (p = 0.11). Urinary NGAL and urinary NGAL/UCr were significantly higher in subjects who met this C_min_ goal (p<.001 for both). KIM-1 and KIM-1/UCr were higher among subjects who attained a C_min_ ≥ 8 µg/mL, although not to a statistically significant degree (p = .07 and p = 0.15, respectively).

## Discussion

In our pilot study, a substantial proportion of critically ill children failed to attain therapeutic targets for FEP with standard dosing early in the course therapy. We evaluated a target of 8 µg/mL, which is the CLSI breakpoint for *Pseudomonas aeruginosa*, a key pathogen often targeted with empiric FEP treatment in the critical care setting.^25^ Only 43% of subjects in our study maintained free FEP concentrations above 8 µg/mL for 100% of their dosing interval, assuming protein binding of 20-39%. Age, eGFR, and urinary biomarker concentrations (NGAL, KIM-1) were correlated with FEP C_min_, with younger age, higher eGFR, and lower urinary biomarkers associated with sub-therapeutic C_min_.

Delayed administration of effective antimicrobials is associated with a nearly five-fold increased risk of death from sepsis in children.^11^ It follows that sub-optimal FEP exposures (i.e. sub-therapeutic concentrations) would also negatively impact outcomes, although this has not been explicitly studied in pediatric patients. In adults with gram-negative bacteremia, achievement of *f*T>MIC of 68-74% was associated with 6-7-fold improved odds of survival, with each 1% increase in *f*T>MIC associated with a 2% increase in survival.^26^ Adults with gram-negative pneumonia who had a C_min_/MIC ≥ 2.1 had a significantly lower risk of clinical failure.^27^ And, adults with serious bacterial infections treated with FEP who had >100% *f*T>MIC had significantly greater clinical cure and bacterial eradication than those with <100% *f*T>MIC.^12^ While a specific FEP target for use in critically ill children has not been established, it is likely that antibiotic exposures are an important part of successful treatment and outcomes.

Therapeutic drug monitoring (TDM) is the practice of measuring drug concentrations to inform dosing in patients. It is typically reserved for drugs with a narrow therapeutic window to optimize safety (primarily), as well as efficacy. β-lactam TDM has become more common among adult patients, particularly those felt to be at-risk for sub-therapeutic concentrations and poor outcomes, such as those who are critically ill.^13^ But, TDM for β-lactam drugs is not routine in pediatric clinical practice. Based on our evaluation, TDM may have utility in critically ill children since a large proportion may fail to attain empiric targets for FEP. This could be achieved by measurement of a single drug concentration as close to the end of the dosing interval as possible, recognizing that mistiming of this single measurement can have a significant impact on interpretability. Alternatively, 2 post-distribution concentrations could be obtained during a single dosing interval and log-linear regression methods used to estimate the elimination rate and C_min_, similar to how we performed our analyses. This approach would allow for more specific dose adjustments and individualized tailoring of therapy compared to trough-only TDM.

A key to TDM is identifying the right patients or patient population in which to perform it. Although FEP neurotoxicity occurs with high C_min_ in adults,^28^ it is a relatively rare occurrence in children. Therefore, it is more important to identify the subset of patients whose concentrations are most likely to fall below the therapeutic range. In our study, younger patients, those with a higher eGFR, and those with low urinary kidney injury biomarkers (i.e. potentially indicating healthier kidneys and better function) were more likely to fail to attain C_min_ targets. Further, each of these factors (age, eGFR, biomarkers) was also significantly correlated with FEP C_min_. Augmented renal clearance (ARC) has been associated with sub-therapeutic antibiotic concentrations in critically ill adults.^29^ But, there has been debate surrounding the specific creatinine clearance (or eGFR) cut point that reflects ARC in children. ARC, defined as a urinary creatinine clearance (CrCl) >130 mL/min/1.73 m^2^, is present in 20-50% of critically ill adults and is associated with subtherapeutic antibiotic concentrations in such patients.^29^ However, there is no standard definition for ARC in children and measurement of urinary CrCl in children is rarely pursued. In our study, ARC was not significantly associated with failure to attain FEP C_min_ ≥ 8 µg/mL, nor was an eGFR >130 mL/min/1.73 m^2^. But, the association between higher eGFR and C_min_ < 8 µg/mL suggests that drug clearance is the major driver of subtherapeutic FEP concentrations.

This is further supported by the association between higher biomarker (NGAL, KIM-1) concentrations and lower C_min_. These two biomarkers have been studied extensively and show to have utility in early detection of acute kidney injury.^30,31^ It follows that higher concentrations, reflective of ongoing injury, would be associated with reduced FEP clearance since FEP is a renally eliminated drug. Because our study was small, we did not seek to define a specific biomarker cut-off that was associated with failure to attain FEP C_min_ targets. But, future large-scale studies may be able to do so. While urinary biomarkers hold promise as noninvasive and sensitive tests for detection of acute kidney injury, they may similarly be useful to identify candidates for β-lactam TDM: those patients at highest risk for therapeutic failure.

It is noteworthy that we used log-linear regression to estimate FEP *f*T>MIC and C_min_. This approach assumes first-order kinetics and is not as robust as some other methods, such as use of Bayesian predictions based on population PK models. Unfortunately, our study was not designed to develop a population PK model given the limited sample size and heterogeneity of the population studied. Instead, we chose our approach to mirror what would most often be done clinically to estimate *f*T>MIC and C_min_ for FEP in children. While Bayesian methods can estimate more accurate PK profiles for individual patients, the use of first order equations is a reasonable and practical alternative to determining target attainment.^32^ Our sampling times were all during the post-distribution phase, samples were collected at steady-state, and the precise timing of sampling was recorded, minimizing bias of this approach. Further, to our knowledge, no currently available Bayesian dosing software programs incorporate pediatric PK models for FEP. Given our findings, larger-scale PK studies of FEP are warranted in critically ill children. Such studies can promote development of population PK models, potentially incorporating novel urinary biomarkers, to support Bayesian dosing and ensure adequacy of FEP dosing and target attainment in this vulnerable population.

There are additional limitations to our study. First, due to our small sample size, we chose to explore only a limited number of factors that we believed were most likely to influence FEP C_min_ (e.g. renal function, severity of illness indicators, dosing information). It is possible that other unexamined factors could be influential. Further, our small sample size precluded pursuit of multivariable analyses or an evaluation of outcomes. Second, our lab measured total FEP concentrations and we estimated the free fraction based on recent published literature.^21^ Previous pediatric studies have utilized a lower fraction of protein binding (20%),^22^ which could impact target attainment estimates. Our sensitivity analyses showed that protein binding had a minimal influence on target attainment estimates in our population. Importantly, the fraction of subjects with a C_min_ ≥8 µg/mL was the same whether 20% or 39% protein binding was assumed. Finally, we did not specify when urine samples should be collected in relation to FEP concentration sampling, aside from urine sampling occurring before completion of PK sampling. We intentionally permitted variability in this approach in hopes that it would mirror clinical care in the PICU setting. While the association between urinary biomarker concentrations and FEP clearance may be stronger if evaluated at a particular interval, this would not be as practical clinically. Larger future studies may better evaluate the optimal timing of urine sample collection to inform FEP TDM.

In conclusion, a significant proportion of critically ill children failed to attain FEP targets in plasma using standard, intermittent dosing. Although all subjects received recommended dosing for treatment of serious infections, less than half attained targeted goal concentrations. Younger age, higher eGFR, and lower urinary NGAL and KIM-1 were associated with failure to attain C_min_ (i.e. 100% *f*T>MIC) targets. Urinary biomarkers may be a non-invasive means to screen for patients at risk for subtherapeutic FEP concentrations.

## Data Availability

All data produced in the present study are available upon reasonable request to the authors.

## Abbreviations

ARC: augmented renal clearance
C_min_: minimum concentration
eGFR: estimated glomerular filtration rate
FEP: cefepime
*f*T>MIC: fraction of time free concentration is above the minimum inhibitory concentration
KIM-1: kidney injury molecule-1
MIC: minimum inhibitory concentration
NGAL: neutrophil gelatinase-associated lipocalin
PELOD-2 score: Pediatric Logistic Organ
Dysfunction-2 score; SCr: serum creatinine

## TRANSPARENCY DECLARATIONS

KJD has received research support from Merck & Co., Inc. unrelated to the current work. AFZ has received research support from Eunice Kennedy Shriver National Institute of Child Health & Human Development (award numbers UG1HD063108, R21HD093369), the U.S. Department of Defense (award number W81XWH-17-1-0668), and Zelda Therapeutics, unrelated to this work. The content is solely the responsibility of the authors and does not necessarily represent the official views of any of the above supporting agencies. All other authors: Nothing to declare.

